# Metabolomics Identifies and Validates Serum Androstenedione as Novel Biomarker for Diagnosing Primary Angle Closure Glaucoma and Predicting the Visual Field Progression

**DOI:** 10.1101/2023.08.08.23293850

**Authors:** Shengjie Li, Jun Ren, Yichao Qiu, Zhendong Jiang, Mingxi Shao, Yingzhu Li, Jianing Wu, Yunxiao Song, Xinghuai Sun, Shunxiang Gao, Wenjun Cao

**Author notes:** These authors contributed equally (Shengjie Li, Shunxiang Gao, Wenjun Cao). Corresponding author: Shengjie Li, Eye & ENT Hospital, Shanghai Medical College, Fudan University, Fenyang Road 83th, Shanghai 200031, China., Shunxiang Gao, Eye & ENT Hospital, Shanghai Medical College, Fudan University, Fenyang Road 83th, Shanghai 200031, China., Wenjun Cao, Eye & ENT Hospital, Shanghai Medical College, Fudan University, Fenyang Road 83th, Shanghai 200031, China.

## Abstract

**Background:** Primary angle closure glaucoma (PACG) is the leading cause of irreversible blindness in Asia, and no reliable, effective diagnostic, and predictive biomarkers are used in clinical routines. A growing body of evidence shows metabolic alterations in patients with glaucoma. we aimed to develop and validate potential metabolite biomarkers to diagnose and predict the visual field progression of PACG.

**Methods:** Here, we used a 5-phases (discovery phase, validation phase 1, validation phase 2, supplementary phase, and cohort phase) multicenter (EENT hospital, Shanghai Xuhui central hospital), cross-sectional, prospective cohort study design to perform widely-targeted metabolomics and chemiluminescence immunoassay to determine candidate biomarkers. Five machine learning (random forest, support vector machine, lasso, K-Nearest neighbor, and Gaussian NB) approaches were used to identify an optimal algorithm. The discrimination ability was evaluated using the area under the receiver operating characteristic curve (AUC). Calibration was assessed by Hosmer-Lemeshow tests and calibration plots.

**Results:** Studied serum samples were collected from 616 participants, and 1464 metabolites were identified. Machine learning algorithm determines that androstenedione exhibited excellent discrimination and acceptable calibration in discriminating PACG across the discovery phase (discovery set 1, areas under the receiver operating characteristic curve [AUC] =1.0 [95%CI, 1.00-1.00]; discovery set, AUC=0.85 [95%CI, 0.80-0.90]) and validation phases (internal validation, AUC=0.86 [95%CI, 0.81-0.91]; external validation, AUC=0.87 [95%CI, 0.80-0.95]).

Androstenedione also exhibited a higher AUC (0.92-0.98) to discriminate the severity of PACG. In the supplemental phase, serum androstenedione levels were consistent with those in aqueous humor (r=0.82, P=0.038) and significantly (P=0.021) decreased after treatment. Further, cohort phase demonstrates that higher baseline androstenedione levels (hazard ratio=2.71 [95% CI: 1.199-6.104], P=0.017) were associated with faster visual field progression.

**Conclusion:** Our study identifies serum androstenedione as a potential biomarker for diagnosing PACG and indicating visual field progression.

## 1. Background

Glaucoma is the most frequent cause of irreversible blindness worldwide^1^, and its prevalence is increasing globally, making it a public health concern^2^. The primary form of glaucoma worldwide is Primary Open-Angle Glaucoma (POAG). However, in East Asian populations, Primary Angle-Closure Glaucoma (PACG) is predominant, affecting 70% of glaucoma patients globally.^3–5^. Hence, early detection of PACG and accurate prediction of visual field (VF) changes can potentially preserve vision and mitigate the risk of PACG advancement. Measurement of intraocular pressure, perimetry, gonioscopy and optical coherence tomography is the main diagnostic testing to assess for glaucoma and to monitor for disease progression^6^. However, the present clinical techniques are inadequate for the early diagnosis and prognosis of VF progression as they depend on specialized eye examination equipment and the expertise of ophthalmologists. Furthermore, patients seldom seek the services of an ophthalmologist until the symptoms worsen or visual acuity significantly deteriorates. Importantly, the He et al.^7^ study taught us that performing laser iridotomy on patients with 180 degrees of angle closure does not prevent PACG, thus gonioscopy may not a good tool to help distinguish PACG from controls. Therefore, the development of a straightforward and dependable biomarker or biomarker panel to facilitate early detection and prognostication of progressive visual field loss in primary angle-closure glaucoma is imperative, rather than relying solely on the expertise of ophthalmologists and specialized equipment.

Multiple etiology and risk factors lead to the onset/development of PACG in humans, which involves a tremendous flow of physiological changes, genetic factors, and metabolic adaptations^8^. Both physiological changes, genetic factors, and metabolic adaptations should lead to profound changes in most metabolic pathways. Small molecule metabolites, as crucial biomarkers of cellular function^9^, which identifies and quantified by metabolomics, are the omics product closest to clinical phenotypes^10^. A burgeoning corpus of evidence indicates metabolic changes in individuals afflicted with glaucoma^11–15^. Nevertheless, extant metabolomics research on glaucoma is constrained by inadequate sample sizes, a dearth of validation sets to corroborate findings, and an absence of specificity analyses. Notably, investigations have yet to comprehensively characterize the serum metabolome in sizable cohorts to identify putative biomarkers capable of distinguishing patients with PACG from healthy controls.

In this study, a cross-sectional and prospective cohort design was employed to systematically profile blood metabolites using widely-targeted metabolomics and chemiluminescence immunoassay in both patients with primary angle-closure glaucoma (PACG) and control individuals. The objectives of the study were to characterize the metabolic profile associated with PACG, identify potential blood diagnostic biomarkers of PACG, assess the specificity of diagnostic biomarkers for PACG of any severity, and verify the biomarkers used to predict the visual field progression of PACG.

## 2. Methods

### 2.1 Participant

From January 2020 to December 2021, newly diagnosed PACG and age-sex-matched controls were recruited from the Eye Center of Fudan University and Shanghai Xuhui Central hospital. Detailed ophthalmic examinations and medical examinations were described in the supplementary material. Approval from the Institutional Review Board/Ethics Committee (2020[2020013]) was obtained from the Ethics Committee of the Eye and ENT Hospital, and the study adhered to the principles of the Declaration of Helsinki. Informed consent was obtained from all subjects.

A glaucoma specialist diagnosed PACG. The diagnostic criteria for PACG were described previously^16–18^ and detailed in the supplementary material. The inclusion and exclusion criteria of patients with PACG were described previously^16–18^ and detailed in the supplementary material. The methods performed for VF analysis were done as previously described^16,18^ and detailed in the supplementary material. Previously described methods^16,18–20^ were performed for the determination of functional PACG VF loss progression according to an event-based analysis modified for Octopus perimetry.

### 2.2 Study design

A cross-sectional, multicenter, prospective cohort study design. In the cross-sectional study, a total of 616 patients and controls were prospectively enrolled from the Eye Center of Fudan University and Shanghai Xuhui Central hospital which was divided into 4 phases (discovery phase [discovery set 1, the discovery set 2], validation phase 1 [external validation], validation phase 2 [internal validation], supplemental phase) from eight districts in China (Figure S1). The 5 phases of the study are independent and are shown in Figure 1. The discovery set 1 was composed of 80 serum samples from PACG patients and 60 samples from controls which was enrolled from Eye Center of Fudan University between January 2020 to June 2020. The discovery set 2 was composed of 100 serum samples from PACG patients and 80 samples from controls which was enrolled from Eye Center of Fudan University between July 2020 to Deccember 2020. The differential metabolites of discovery set 1 and discovery set 2 were obtained by widely-targeted metabolomics. The intersection of the differential metabolites of the two discovery sets was used as a candidate biomarker. For validation phase 1, serum samples from 70 PACG patients and 50 controls were collected which was enrolled from Shanghai Xuhui Central hospital between January 2020 to December 2020. Candidate biomarkers were validated in validation phase 1 using widely-targeted metabolomics, and potential biomarkers were obtained. For validation phase 2, serum samples from 98 PACG patients and 78 controls were collected which was enrolled from Eye Center of Fudan University between January 2021 to December 2021. Potential biomarkers were validated in validation phase 2 using chemiluminescence methods. For the supplemental phase, we used widely-targeted metabolomics to investigate whether the same potential biomarkers were present in PACG patients’ aqueous humor. All the clinical characteristics (Age, sex, BMI, hypercholesterolemia, hypertension, diabetes, smoking, drinking) were matched between PACG and controls in the 4 phases.

**Figure 1.**
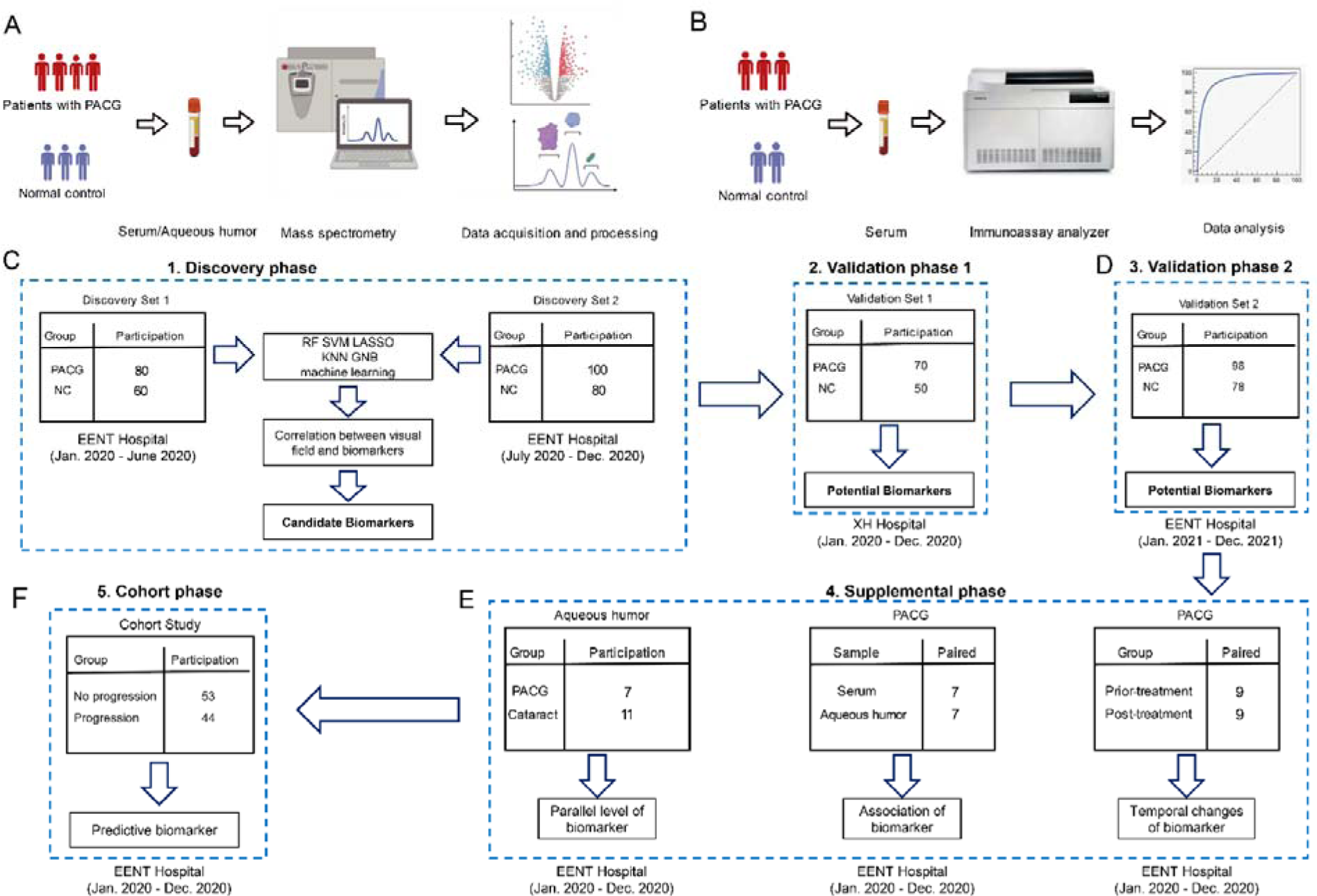
Study design and workflow. A 5-phase study (discovery phase [discovery set 1, the discovery set 2], validation phase 1, validation phase 2, supplemental phase, and cohort phase) design. A: The workflow of the discovery phase, validation phase 1, supplemental phase, and cohort phase were analyzed by liquid chromatography-mass spectrometry (LC-MS) for untargeted metabolomics. B: The workflow of validation phase 2 was analyzed chemiluminescence immunoassay for targeted detection. C: A total of 440 patients and controls were recruited and assigned to discovery set 1 (n = 140), the discovery set 2 (n = 180), and validation set 1 (n = 120). The biomarker signature was identified on the metabolomic data from the discovery phase, comparing primary angle closure glaucoma (PACG) with control patients. These data were used as a discovery set for the algorithm. D: Validation phase 2 (n = 176) was included as the second validation cohort. E: Three measurements were performed in the supplemental phase. F: Cohort phase were performed to validate the predictive value of biomarker (n = 97).

In the prospective cohort study, 98 newly diagnosed PACG patients were included from the Eye Center of Fudan University between January 2020 and December 2020. All participants visited once every six months to allow regular assessment of PACG disease progression (the minimum follow-up period was set to 24 months). Serum samples from 98 PACG patients were collected and measured using chemiluminescence methods. Detailed information of patients follow up were described as previously^16,18^.

### 2.3 Sample preparation

#### 2.3.1 Sample collection

The blood was collected prior to the medical or surgical treatment. Briefly, subjects were asked to ensure about eight-ten hours of fasting before sampling. Venous blood samples were collected in heparinized tubes between 7:00-9:30 am. The clinical laboratory obtained the sample at about 10:00 am and centrifuged them at 3000rpm for 10min. Then serum was collected into a sterilized cryotube and immediately stored at –80□ for metabolomic analysis.

The acquisition of aqueous humor was described previously^13^. Aqueous humor (AH) was collected by skilled ophthalmologists during the surgical treatment of PACG and cataract patients. AH was obtained at the start of the procedure using a fine-bore needle during a corneal paracentesis. Following that, AH samples were quickly transferred to sterile cryotubes and kept at –80 °C for metabolomic analysis.

#### 2.3.2 Sample preprocessing

The same internal standard was added to each sample during metabolite extraction (L-2-chlorophenylalanine, 4-Fluoro-L-α-phenylglycine, [2H5]-Kynurenic Acid, [2H5]-Phenoxy acetic Acid, Indole-3-butyric-2,2-d2 Acid, LysoPC 19:0, DL-3-Indole-lacticacid). Before extraction, the serum was thawed on ice, vortex for 10s and mixed well. 300µL of pure cold methanol was added to 50µL of serum, swirled for 3min, and centrifuged at 12000 rpm for 10min at 4°C. After centrifugation, transfer the supernatant into a new centrifuge tube and place it in a –20 [refrigerator for 30 minutes. The sample was thawed on ice and centrifuged at 12000r/min for 3 minutes— Transfer 180µL of the supernatant to the injection vial for mass spectrometry analysis. Quality control (QC) samples are generated by pooling all the serum samples to monitor the retention time and signal intensity consistency. Equal volumes (10 µL) of all serum samples were combined to generate quality control (QC) samples, which were employed to monitor the repeatability of the analysis. During the mass spectrometry analysis, one QC sample was included in every ten samples to ensure the consistency of the analytical process(sheet1). The PACG and control groups’ serum samples were randomly arranged and analyzed. The laboratory conducting the metabolomics measurements was blinded to the samples’ case/control/QC status.

### 2.4 Analytical methods ^21^

#### 2.4.1 Untargeted metabolomics analysis

Ultra Performance Liquid Chromatography (UPLC) (ExionLC AD, AB SCIEX) separation was performed using Waters ACQUITY HSS T3 (2.1×100mm, 1.8um). The oven temperature was set to 40[, and the sample injection volume was 5 µL. Metabolites were eluted from the column at a flow rate of 0.35 mL/min. Mobile phases for UPLC consisted of 0.1% acetic acid in water (phase A) and 0.1% acetic acid in acetonitrile (phase B). The following gradient elution program was employed: 0-10min: linear gradient from 5% to 90% B; 10-11min: 90% B; 11-11.1min: linear gradient from 90% to 5% B; 11.1-14min: 5% B. Metabolic extracts mixture (QC sample) were analyzed by the triple time of flight (TOF) mass spectrometer (TripleTOF 6600, AB SCIEX) in both positive and negative ionization modes. The scan range was 50-1,000 m/z. Electrospray ionization (ESI) source conditions were set as follows: ion spray voltage (IS) 5500V (positive), –4500V (negative); ion source gas I (GSI), gas II (GASII), curtain gas (CUR) was set at 50,50, and 25 psi, respectively; collision energy (CE) 30V.

#### 2.4.2 Widely targeted detection conditions

UPLC (ExionLC AD, AB SCIEX) separation was performed using Waters ACQUITY UPLC C18(2.1×100mm, 1.8um). The oven temperature was set to 40[, and the sample injection volume was 2 µL. Metabolites were eluted from the column at a flow rate of 0.35 mL/min. Mobile phases for UPLC consisted of 0.1% acetic acid in water (phase A) and 0.1% acetic acid in acetonitrile (phase B). The following gradient elution program was employed: 0-11min: linear gradient from 5% to 90% B; 11-12min: 90% B; 12-12.1min: linear gradient from 90% to 5% B; 12.1-14min: 5% B. Metabolic extracts of each sample were analyzed by the triple quadrupole-linear ion trap mass spectrometer (QTRAP 6500, AB SCIEX) in both positive and negative ionization modes. ESI source conditions were set as follows: ion spray voltage (IS) 5500V (positive), –4500V (negative); ion source gas I (GSI), gas II (GASII), curtain gas (CUR) was set at 50,50, and 25 psi, respectively. Each ion pair is scanned for detection based on optimized voltage and CE.

#### 2.4.3 Metabolite profiling

Mixed samples (QC sample) were made and tested by AB Triple TOF 6600 mass spectrometer, the metabolites identified base on public database including Metware public database (Metlin, HMDB, KEGG), and MetDNA. The detected metabolites of QC sample (metabolites with a total score > 0.5) add Metware in-house database to be a new whole database. MRM was used for each samples to determine the final ion pair and other information. Based on the new database, Q-trap 6500 was used to quantify all samples accurately. The workflow of mass spectrometry is detailed in Figure S2.

#### 2.4.4 Data processing

All LC-MS data were processed using Analyst 1.6.3 for imputing raw, peak picking, alignment, normalization, and to produce peak intensities for retention time and m/z data pages. The features were selected based on their CV with QC samples. Features with CVs of more than 15% were eliminated.

#### 2.4.5 Chemiluminescence immunoassay

Serum levels of androstenedione were measured using a commercially available kit (Snibe Diagnostics, Shenzhen, China) and were determined using the chemiluminescent immunoassay method by Roche Cobase e 601 (Germany).

### 2.5 Sample size and missing value

In the cross-sectional study, to calculate the minimum total sample size, we used an open-source calculator based on the methods described by Obuchowski et al.^22^ and Li et al.^23^. The input parameters were specificity = 0.9 (allowable error = 0.05), sensitivity = 0.9 (allowable error = 0.05), α = 0.05 (2-tailed). Based on this calculation, the minimum sample size required for the new biomarker was 98 per phase.

An unreliable conclusion would result from missing data, which would introduce bias. Thus no patients with missing data were included in this study. In this study, a total of 48 participants (lack of VF value = 18, lack of medication history record = 12, lack of intraocular pressure [IOP] value = 10, loss to follow-up = 8) were excluded due to with missing data.

### 2.6 Statistical analysis

Normality was assessed using the Shapiro–Wilk W-test. Independent student’s t-test, Kruskal-Wallis test, one-way ANOVA, Wilcox test, and chi-square tests were used when appropriate. Results are presented as frequency and percentage for categorical variables, mean ± SD for normally distributed continuous variables, and median (interquartile range) for not normally distributed continuous variables. The Spearman correlation test was used to determine the significance of the correlations between the variables.

Diagnostic efficiency was evaluated using receiver operating characteristic (ROC) curves. Five machine learning (random forest, support vector machine, lasso, K-nearest neighbor [KNN], and Gaussian Naive Bayes [NB]) approaches were used to identify an optimal algorithm. The Youden index maximizing sensitivity plus specificity is applied to determine the best cutoff value, sensitivity, specificity, accuracy, positive predictive value (PPV), and negative predictive value (NPV) were also calculated. Hosmer-Lemeshow tests were used to assess the goodness of fit. The calibration of the biomarker was assessed by computing the calibration plot. Hanley-McNeil method was used to compare these AUCs.

Unsupervised principal component analysis (PCA) was performed by statistics function prcomp withe R. The data was unit variance scaled before unsupervised PCA. Supervised orthogonal projections to latent structures-discriminate analysis (OPLS-DA) were applied to obtain a higher level of group separation and a better understanding of variables responsible for classification. Heatmaps of samples and metabolites were carried out by R package (ComplexHeatmap; heatmap). In order to evaluate the binding mode of metabolites and proteins, Autodock Vina v.1.2.2 was used to analyze molecular docking. Fold change= the characteristic peak area of metabolites in the PACG group / the control group.

Identified metabolites were annotated using the KEGG Compound database (http://www.kegg.jp/kegg/compound/). Then annotated metabolites were mapped to the KEGG Pathway database (http://www.kegg.jp/kegg/pathway.html). Significantly enriched pathways were identified with a hypergeometric test’s p-value for metabolites.

Cox proportional hazards analysis was also carried out to investigate the relationship between baseline androstenedione levels and VF progression loss. The HRs and baseline characteristics that would assist in categorizing participants into the non-progressing PACG group over the follow-up period were determined using Cox proportional hazards models. Kaplan-Meier plots were used to study the survival results, and the log-rank test was applied to see whether there were any differences between the produced plots.

All statistical analyses were performed using R programming language and SPSS 13.0 (SPSS Inc., Chicago, IL, USA), a list of statistical approaches and packages detailed in table S1. P-values less than 5% were considered statistically significant.

## 3. Results

### 3.1 Metabolomic analyses in samples from PACG patients and normal controls

The design of this study is depicted in Figure 1, while the clinical features of all participants in the four phases of the study are presented in Tables 1 (Phases 1-3) and S2-S3 (Phase 4). All clinical characteristics, including age, sex, BMI, hypercholesterolemia, hypertension, diabetes, smoking, and drinking, were carefully matched between the PACG and normal control groups across all four phases.

**Table 1.**
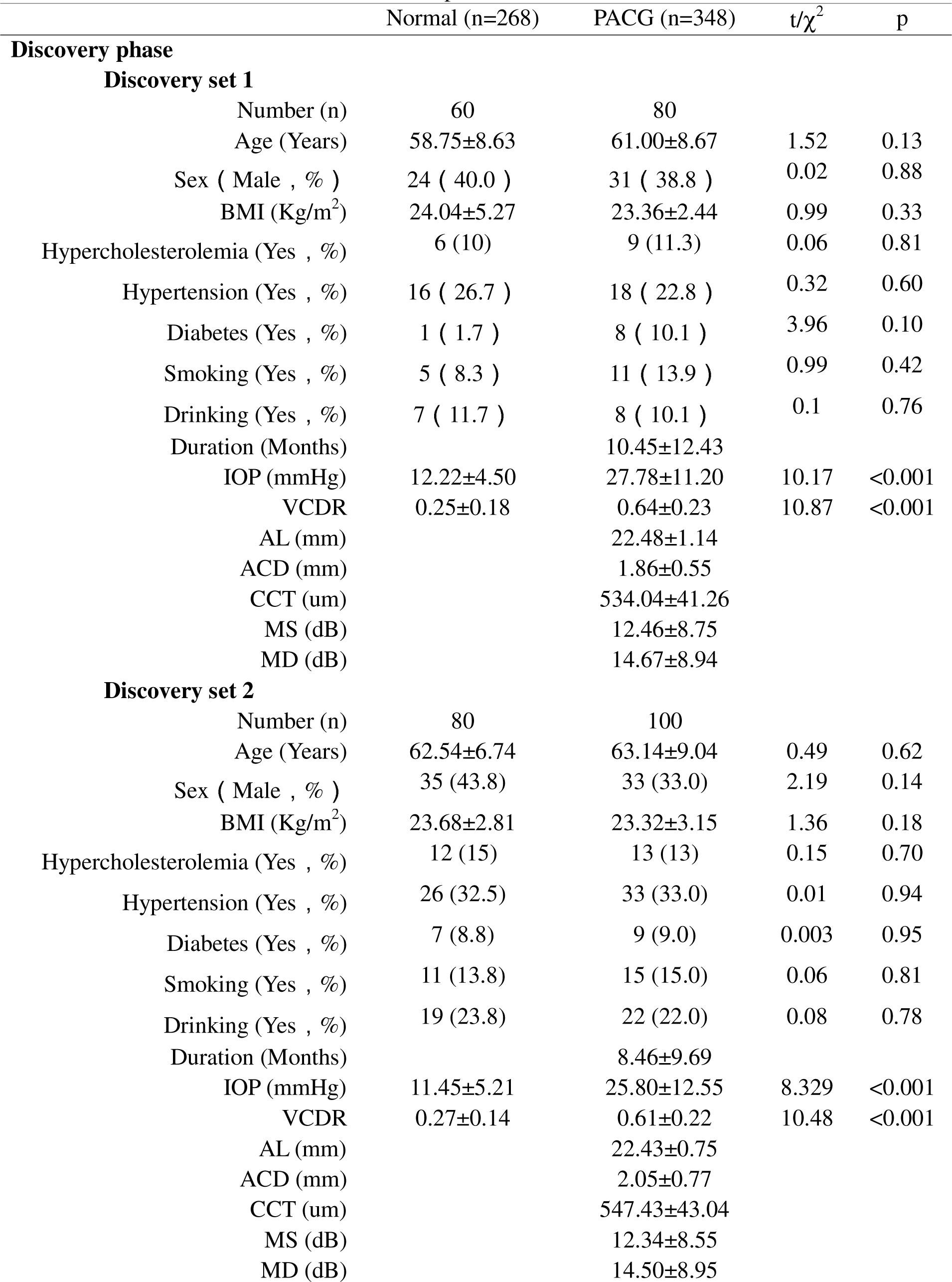
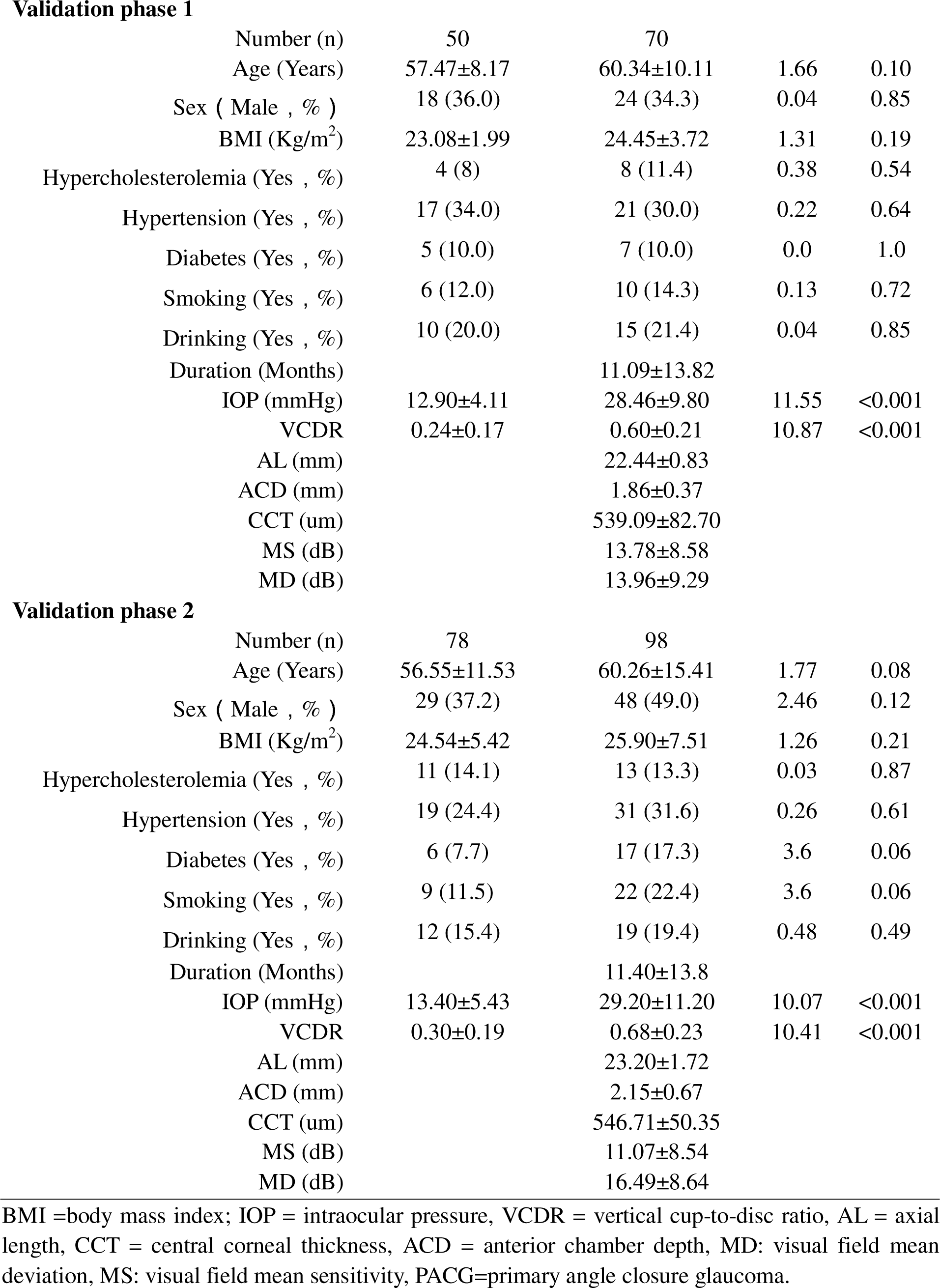
The clinical and demographic characteristics of all subjects in the discovery and validation phases.

| | Normal (n=268) | PACG (n=348) | t/$\chi^2$ | p |
| --- | --- | --- | --- | --- |
| **Discovery phase** |  |  |  |  |
| **Discovery set 1** |  |  |  |  |
| Number (n) | 60 | 80 |  |  |
| Age (Years) | 58.75±8.63 | 61.00±8.67 | 1.52 | 0.13 |
| Sex ( Male , % ) | 24 ( 40.0 ) | 31 ( 38.8 ) | 0.02 | 0.88 |
| BMI (Kg/m<sup>2</sup>) | 24.04±5.27 | 23.36±2.44 | 0.99 | 0.33 |
| Hypercholesterolemia (Yes , %) | 6 (10) | 9 (11.3) | 0.06 | 0.81 |
| Hypertension (Yes , %) | 16 ( 26.7 ) | 18 ( 22.8 ) | 0.32 | 0.60 |
| Diabetes (Yes , %) | 1 ( 1.7 ) | 8 ( 10.1 ) | 3.96 | 0.10 |
| Smoking (Yes , %) | 5 ( 8.3 ) | 11 ( 13.9 ) | 0.99 | 0.42 |
| Drinking (Yes , %) | 7 ( 11.7 ) | 8 ( 10.1 ) | 0.1 | 0.76 |
| Duration (Months) |  | 10.45±12.43 |  |  |
| IOP (mmHg) | 12.22±4.50 | 27.78±11.20 | 10.17 | <0.001 |
| VCDR | 0.25±0.18 | 0.64±0.23 | 10.87 | <0.001 |
| AL (mm) |  | 22.48±1.14 |  |  |
| ACD (mm) |  | 1.86±0.55 |  |  |
| CCT (um) |  | 534.04±41.26 |  |  |
| MS (dB) |  | 12.46±8.75 |  |  |
| MD (dB) |  | 14.67±8.94 |  |  |
| **Discovery set 2** |  |  |  |  |
| Number (n) | 80 | 100 |  |  |
| Age (Years) | 62.54±6.74 | 63.14±9.04 | 0.49 | 0.62 |
| Sex ( Male , % ) | 35 (43.8) | 33 (33.0) | 2.19 | 0.14 |
| BMI (Kg/m<sup>2</sup>) | 23.68±2.81 | 23.32±3.15 | 1.36 | 0.18 |
| Hypercholesterolemia (Yes , %) | 12 (15) | 13 (13) | 0.15 | 0.70 |
| Hypertension (Yes , %) | 26 (32.5) | 33 (33.0) | 0.01 | 0.94 |
| Diabetes (Yes , %) | 7 (8.8) | 9 (9.0) | 0.003 | 0.95 |
| Smoking (Yes , %) | 11 (13.8) | 15 (15.0) | 0.06 | 0.81 |
| Drinking (Yes , %) | 19 (23.8) | 22 (22.0) | 0.08 | 0.78 |
| Duration (Months) |  | 8.46±9.69 |  |  |
| IOP (mmHg) | 11.45±5.21 | 25.80±12.55 | 8.329 | <0.001 |
| VCDR | 0.27±0.14 | 0.61±0.22 | 10.48 | <0.001 |
| AL (mm) |  | 22.43±0.75 |  |  |
| ACD (mm) |  | 2.05±0.77 |  |  |
| CCT (um) |  | 547.43±43.04 |  |  |
| MS (dB) |  | 12.34±8.55 |  |  |
| MD (dB) |  | 14.50±8.95 |  |  |
| **Validation phase 1** |  |  |  |  |
| Number (n) | 50 | 70 |  |  |
| Age (Years) | 57.47±8.17 | 60.34±10.11 | 1.66 | 0.10 |
| Sex ( Male , % ) | 18 (36.0) | 24 (34.3) | 0.04 | 0.85 |
| BMI (Kg/m<sup>2</sup>) | 23.08±1.99 | 24.45±3.72 | 1.31 | 0.19 |
| Hypercholesterolemia (Yes , %) | 4 (8) | 8 (11.4) | 0.38 | 0.54 |
| Hypertension (Yes , %) | 17 (34.0) | 21 (30.0) | 0.22 | 0.64 |
| Diabetes (Yes , %) | 5 (10.0) | 7 (10.0) | 0.0 | 1.0 |
| Smoking (Yes , %) | 6 (12.0) | 10 (14.3) | 0.13 | 0.72 |
| Drinking (Yes , %) | 10 (20.0) | 15 (21.4) | 0.04 | 0.85 |
| Duration (Months) |  | 11.09±13.82 |  |  |
| IOP (mmHg) | 12.90±4.11 | 28.46±9.80 | 11.55 | <0.001 |
| VCDR | 0.24±0.17 | 0.60±0.21 | 10.87 | <0.001 |
| AL (mm) |  | 22.44±0.83 |  |  |
| ACD (mm) |  | 1.86±0.37 |  |  |
| CCT (um) |  | 539.09±82.70 |  |  |
| MS (dB) |  | 13.78±8.58 |  |  |
| MD (dB) |  | 13.96±9.29 |  |  |
| **Validation phase 2** |  |  |  |  |
| Number (n) | 78 | 98 |  |  |
| Age (Years) | 56.55±11.53 | 60.26±15.41 | 1.77 | 0.08 |
| Sex ( Male , % ) | 29 (37.2) | 48 (49.0) | 2.46 | 0.12 |
| BMI (Kg/m<sup>2</sup>) | 24.54±5.42 | 25.90±7.51 | 1.26 | 0.21 |
| Hypercholesterolemia (Yes , %) | 11 (14.1) | 13 (13.3) | 0.03 | 0.87 |
| Hypertension (Yes , %) | 19 (24.4) | 31 (31.6) | 0.26 | 0.61 |
| Diabetes (Yes , %) | 6 (7.7) | 17 (17.3) | 3.6 | 0.06 |
| Smoking (Yes , %) | 9 (11.5) | 22 (22.4) | 3.6 | 0.06 |
| Drinking (Yes , %) | 12 (15.4) | 19 (19.4) | 0.48 | 0.49 |
| Duration (Months) |  | 11.40±13.8 |  |  |
| IOP (mmHg) | 13.40±5.43 | 29.20±11.20 | 10.07 | <0.001 |
| VCDR | 0.30±0.19 | 0.68±0.23 | 10.41 | <0.001 |
| AL (mm) |  | 23.20±1.72 |  |  |
| ACD (mm) |  | 2.15±0.67 |  |  |
| CCT (um) |  | 546.71±50.35 |  |  |
| MS (dB) |  | 11.07±8.54 |  |  |
| MD (dB) |  | 16.49±8.64 |  |  |

BMI =body mass index; IOP = intraocular pressure, VCDR = vertical cup-to-disc ratio, AL = axial length, CCT = central corneal thickness, ACD = anterior chamber depth, MD: visual field mean deviation, MS: visual field mean sensitivity, PACG=primary angle closure glaucoma.

Following rigorous quality control, data filtering, and normalization procedures, a total of 1464 metabolites were identified across the various samples (N = 440). During the discovery phase, the OPLS-DA analysis revealed notable distinctions between participants with primary angle-closure glaucoma (PACG) and those without the condition (Figure 2A). Volcano plots (Figure 2B) were generated using metabolites exhibiting a false discovery rate [FDR] of less than 0.1 and fold changes greater than 1.15 or less than 0.85. In discovery sets 1 and 2, 268 metabolites (21.5%) and 117 metabolites (9.6%), respectively, were found to be significantly altered between PACG and normal subjects.

**Figure 2.**
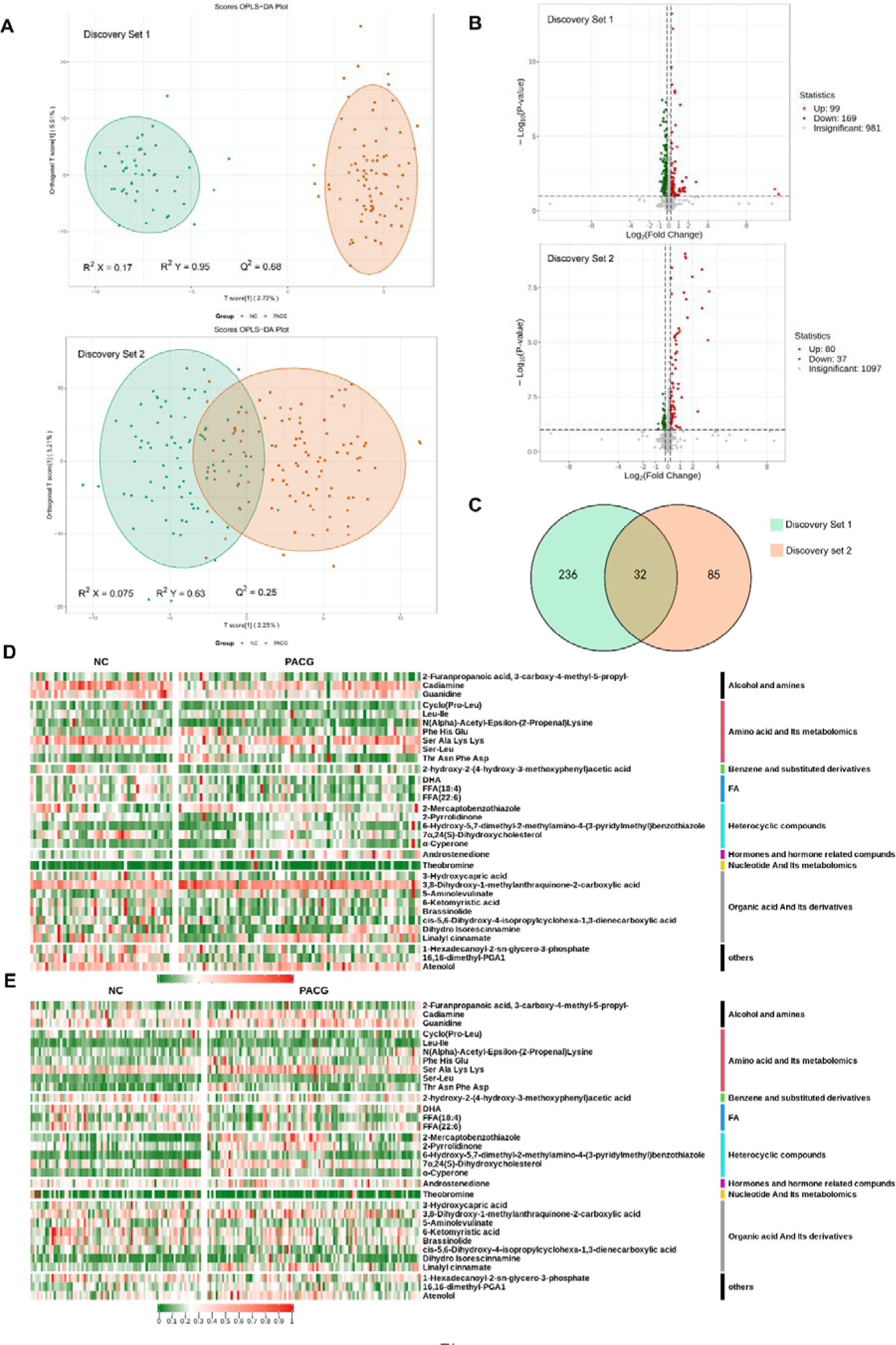
Metabolic profiles discriminate participants with primary angle closure glaucoma (PACG) from normal controls (NC). A: Orthogonal projection to latent structure-discriminant analysis (OPLS-DA) score plot of the comparison between the PACG and NC groups in the discovery phase (discovery set 1 and discovery set 2). Samples in the encircled areas are within the 95% confidence interval. B: Volcano plot of differential metabolites. Metabolites with a fold change of <0.85 and a false discovery rate (FDR) of <0.1 were considered significantly down-regulated. Metabolites with a fold change of >1.15 and an FDR of <0.1 were considered significantly up-regulated. Changes in other metabolites were not significant (insignificant). C: Venn diagram displaying the 32 differential metabolites that were altered as biomarker candidates from the two comparisons in the discovery phase. D: Heatmap of differential metabolites in the discovery set 1 (Data were normalized to min-max). E: Heatmap of differential metabolites in the discovery set 2 (Data were normalized to min-max). FA: fatty acid.

The Venn diagram depicted in Figure 2C illustrates the presence of 32 metabolites that were found to be common in both discovery set 1 (as presented in table S4) and discovery set 2 (as presented in table S5). The parameters used for detecting these 32 differential metabolites are detailed in Table S6. Subsequently, these differential metabolites were utilized for conducting clustering analysis, which is visually represented in the form of a heatmap for both discovery set 1 (Figure 2D) and discovery set 2 (Figure 2E). These 32 differential metabolites were mainly related to alcohol and amines, amino acid and its metabolomics, benzene and substituted derivatives, fatty acid, heterocyclic compounds, hormones, and hormone-related compounds, nucleotide and its metabolomics, organic acid and its derivatives class (Figure 2D, 2E).

### 3.2 The blood differential metabolite discriminates PACG from normal

Can these metabolites in blood differentials be considered as potential biomarkers for PACG? In order to investigate this, we computed the area under the curve (AUC) for each of the 32 differential metabolites to evaluate their discriminatory capacity in distinguishing PACG from healthy individuals using five machine learning techniques (random forest, support vector machine, lasso, KNN, and Gaussian NB). Among these approaches, KNN demonstrated the highest performance in identifying PACG from normal controls. Subsequently, a 2-column heatmap was generated to display the resulting area under the curve (AUC) values for discovery set 1 and discovery set 2, respectively, as depicted in Figure 3A. The receiver operating characteristic (ROC) analysis of 32 metabolites revealed AUC values ranging from 0.74 to 1.0 in discovery set 1 and 0.72 to 1.0 in discovery set 2 for distinguishing PACG from normal subjects. The eigenmetabolite of the 32-metabolite cluster between PACG and normal subjects is illustrated in Figure 4B and 4C. The discriminatory accuracy of the metabolite in differentiating PACG from normal subjects using random forest, support vector machine, lasso, and Gaussian NB is presented in Figure S3.

**Figure 3.**
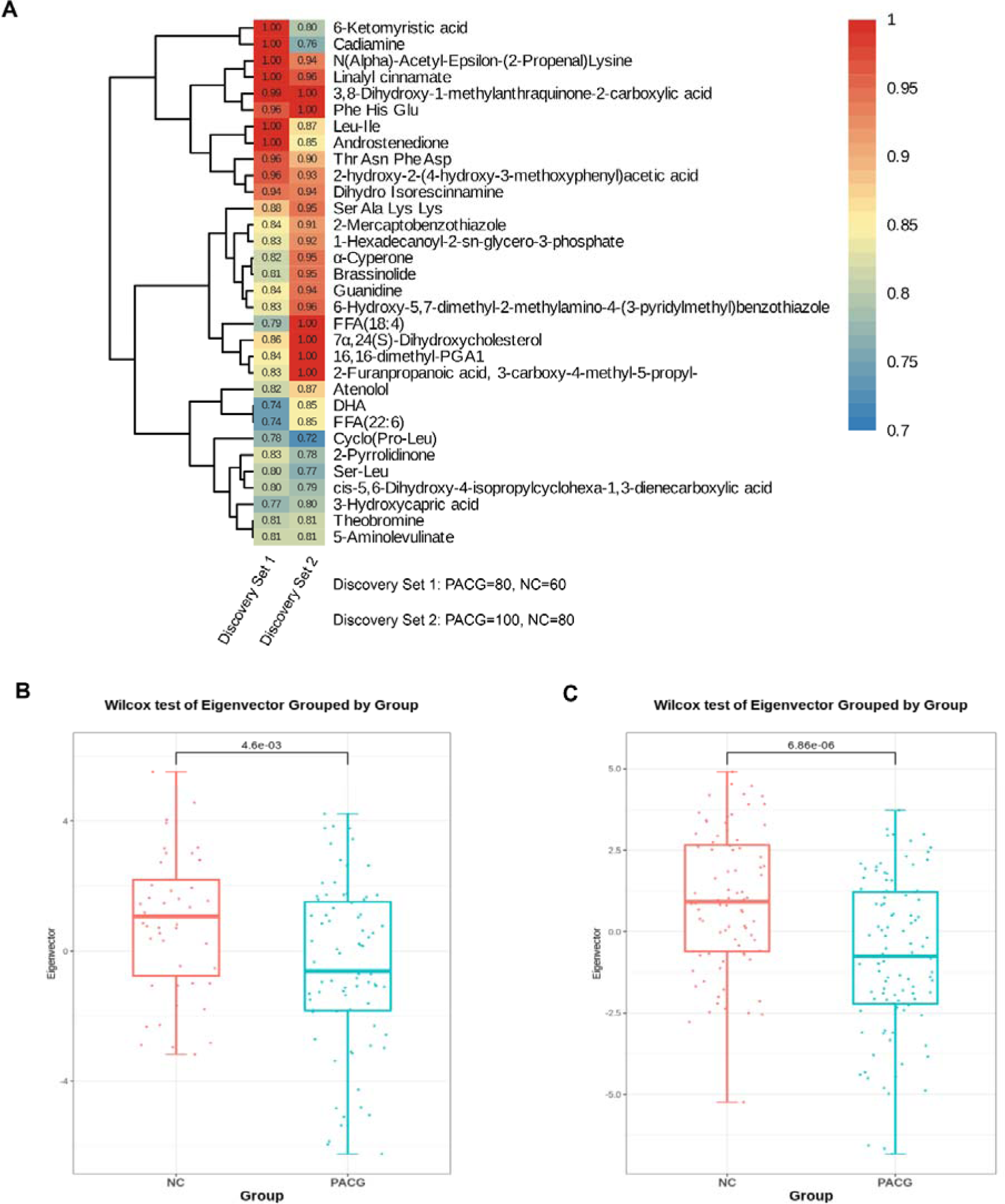
Identification of a unique PACG-associated blood metabolite fingerprint and its behavior in the discovery phase. A: Heatmap of the area under the receiver-operating-characteristic curve assessing the discriminating accuracy of each of the 32 metabolites in differentiating PACG from normal control in the discovery set 1 and discovery set 2. B: The eigenmetabolite of the 32-metabolite cluster between primary angle closure glaucoma (PACG) and control patients in the discovery set 1. C: The eigenmetabolite of the 32-metabolite cluster between primary angle closure glaucoma (PACG) and control patients in the discovery set 2. Wilcox test was used.

**Figure 4.**
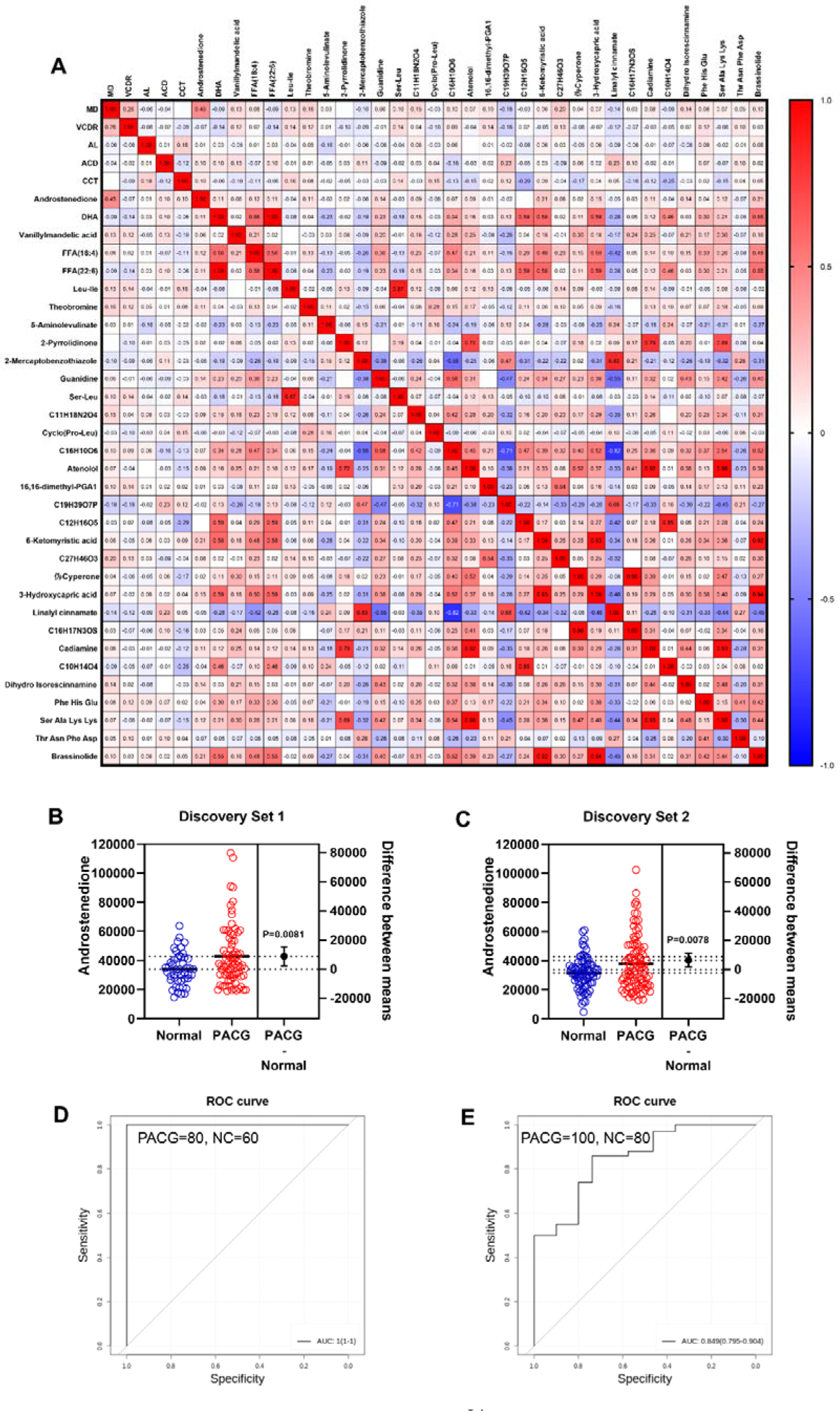
Biomarker discovery discriminates primary angle closure glaucoma (PACG) from normal in the discovery phase. A: A heatmap of correlation analysis between ocular clinical characteristics and 32 potential biomarkers in the discovery phase in PACG subjects. B: The serum level of androstenedione between PACG(42852±20767) and normal(33987±11113) group in the discovery set 1 (Unit for y-axis is peak areas). C: The serum level of androstenedione between PACG and normal group in the discovery set 2 (Unit for y-axis is peak areas). D: Receiver operating characteristic curves of androstenedione to discriminate PACG from normal in the discovery set 1. E: Receiver operating characteristic curves of androstenedione to discriminate PACG from normal in the discovery set 2. Independent student’s t-test was used.

The present study reveals that the eigenmetabolite levels were markedly elevated (P<0.001) in the normal group compared to the PACG group, as evidenced by the results obtained from discovery set 1 (Figure 3B) and discovery set 2 (Figure 3C). These findings provide compelling evidence that the heightened risk of PACG is linked to a robust blood metabolite signature.

### 3.3 Biomarker discovery to discriminates PACG from normal

Figure 3A illustrates that 32 metabolites were verified during the discovery phase and were subsequently identified as potential biomarkers. Figure 4A depicts a correlation analysis between ocular clinical characteristics and the 32 potential biomarkers discovered during the PACG discovery phase. Notably, a significant positive correlation was observed between androstenedione and the mean deviation of the VF (MD) (r=0.45, P<0.001). This correlation was also observed in discovery set 1 (Figure S3, r=0.37, P<0.001) and discovery set 2 (Figure S4, r=0.50, P<0.001), respectively. A statistically insignificant (P>0.05) correlation was observed between ocular clinical characteristics and the remaining 31 potential biomarkers, as depicted in Figure 3A, Figure S3, and Figure S4. Notably, the level of androstenedione was found to be significantly higher in PACG patients than in normal subjects in both discovery set 1 (Figure 4B, P=0.0081, Normal:33987 ± 11113, PACG:42852 ± 20767) and discovery set 2 (Figure 4C, P=0.0078, Normal:31559 ± 10975, PACG:37934 ± 18529). Additionally, high levels of androstenedione were identified as an independent risk factor for PACG, as shown in Table S7. Figure S5 (Discovery set 1) and Figure S6 (Discovery set 2) illustrate the levels of the remaining 31 metabolites between PACG patients and normal subjects. Based on the preceding analysis, androstenedione was chosen using KNN due to its ability to differentiate between PACG and normal subjects. In discovery set 1, the AUC for PACG versus control was 1.0 (95% CI, 1.0 to 1.0), as shown in Figure 4D and Table S8. Similarly, in discovery set 2, PACG was identified with an AUC of 0.85 (95% CI, 0.80 to 0.90) when compared to control individuals, as depicted in Figure 4E and Table S8.

### 3.4 Biomarker validation in two independent validation phases

During the validation phases, Androstenedione was evaluated as a biomarker signature. The samples from validation phase 1 were subjected to LC-MS analysis for widely-targeted metabolomics, which revealed significant differences between PACG and normal participants, as demonstrated by the OPLS-DA analysis (Figure S7). In validation phase 2, a chemiluminescence immunoassay method was developed to enable precise quantification of serum androstenedione levels in a convenient and rapid manner.

During validation phase 1 and 2, the level of androstenedione was found to be significantly higher in individuals with PACG compared to normal subjects (Figure 5A, P=0.0042, Normal:60737 ± 28078, PACG:82394 ± 33994; Figure 5B, P=0.0034, Normal:1.552 ± 0.489, PACG:1.825 ± 0.6876). The performance of androstenedione was evaluated using the AUC in ROC analysis. The AUC for PACG versus control was 0.87 (95% CI, 0.80 to 0.95) in validation phase 1, as depicted in Figure 5C and Table S8. In validation phase 2, a consistent performance of androstenedione (AUC, 0.86, 95% CI, 0.81 to 0.91) was observed (Figure 5D, Table S8).

**Figure 5.**
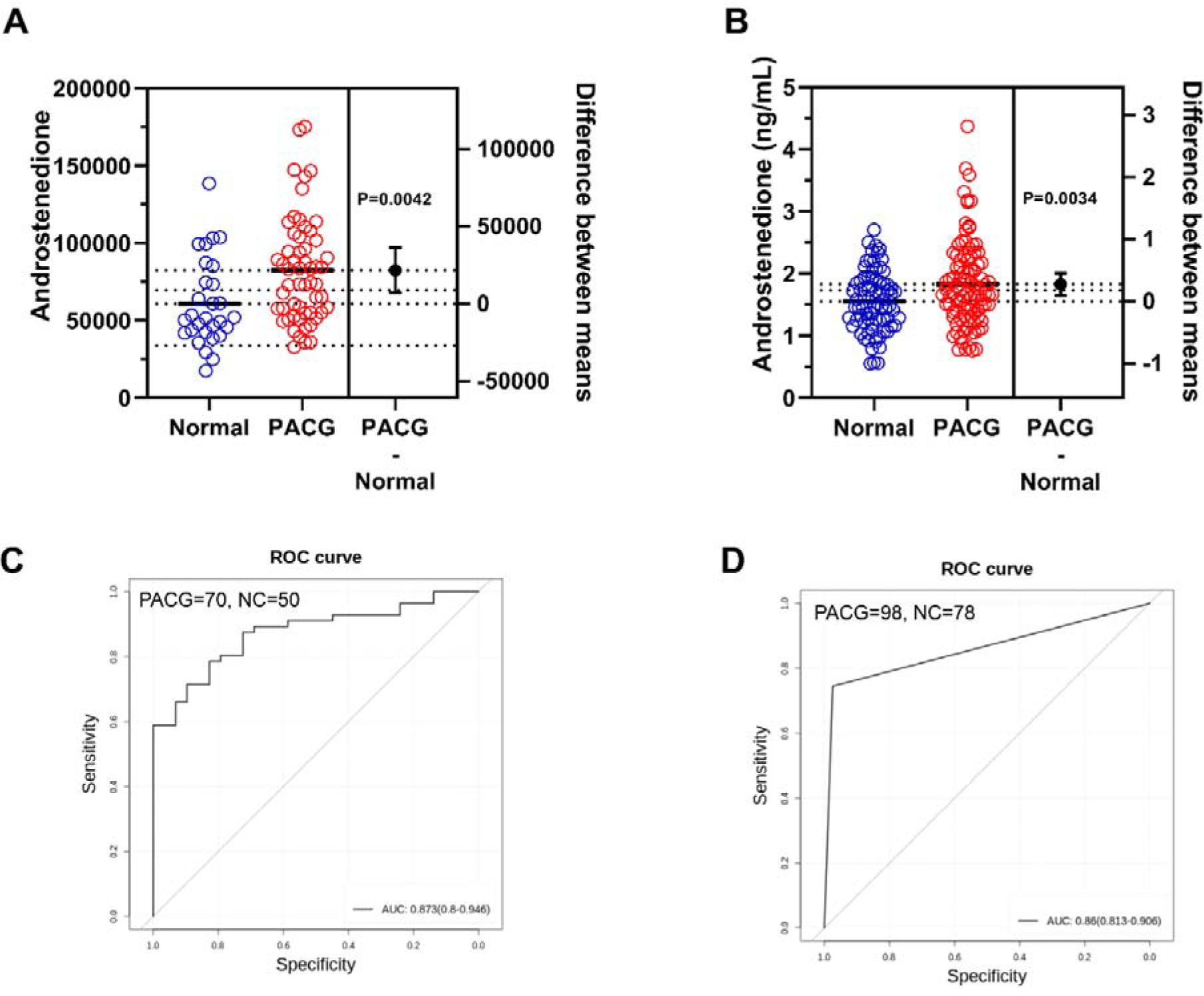
Biomarker validation in two independent validation phases to discriminate primary angle closure glaucoma (PACG) from normal. A: The serum level of androstenedione between PACG and normal group in validation phase 1 (Unit for y-axis is peak areas). B: The serum level of androstenedione between PACG and normal group in validation phase 2 (Unit for y-axis is peak areas). C: Receiver operating characteristic curves of androstenedione to discriminate PACG from normal in validation phase 1. D: Receiver operating characteristic curves of androstenedione to discriminate PACG from normal in validation phase 2. Independent student’s t-test was used.

### 3.5 Biomarker validation in male and female subgroups

Validation of androstenedione as a biomarker for PACG necessitates consideration of sex, as males exhibit 5-10 times higher levels of this hormone than females. Accordingly, the subjects were stratified into male and female subgroups.

Figure S8 demonstrates that in the discovery phase, the AUC for PACG compared to control was 1.0 (95% CI, 1.0 to 1.0) in the male subgroup (Figure S8A), and 0.91 (95% CI, 0.85 to 0.98) in the female subgroup (Figure S8B). In validation phase 1, the AUC for PACG versus control was 0.83 (95% CI, 0.73 to 0.92) in the male subgroup (Figure S8C), and 1.0 (95% CI, 1.0 to 1.0) in the female subgroup (Figure S8D). In validation phase 2, the AUC for PACG versus control was 1.0 (95% CI, 1.0 to 1.0) in the male subgroup (Figure S8E), and 0.88 (95% CI, 0.79 to 0.98) in the female subgroup (Figure S8F).

### 3.6 Androstenedione associates with severity of PACG

The present study aimed to examine the potential association between serum androstenedione levels and the clinical severity of PACG. PACG severity was categorized into mild (MD≦6), moderate (6-12), and severe (MD >12) based on the MD value. The results from both discovery set 1 (Figure S9A, Mild:32600 ± 17011, Moderate:33215 ± 17855, Severe:46060 ± 21789) and discovery set 2 (Figure S9B, Mild:27866 ± 19873, Moderate:27057 ± 13166, Severe:43972 ± 19234) indicated that the mean serum androstenedione levels were significantly higher in the severe PACG group compared to the moderate and mild PACG groups (P<0.001). These findings were further validated in both validation phase 1 (Figure S9C, Mild:75726 ± 45719, Moderate:65798 ± 30610, Severe:94348 ± 30858) and validation phase 2 (Figure S9D, Mild:1.121 ± 0.3143 ng/ml, Moderate:1.461 ± 0.4391 ng/ml, Severe:2.147 ± 0.6476 ng/ml).

A correlation analysis was conducted to examine the relationship between MD and androstenedione. Notably, a statistically significant positive correlation was found between MD and androstenedione in both discovery set 1 (Figure S3, r=0.37, P<0.001) and discovery set 2 (Figure S4, r=0.50, P<0.001). This finding was further confirmed in validation phase 2 (Figure S9E, r=0.61, P<0.001). Subsequently, the diagnostic potential of androstenedione in distinguishing the severity of PACG was investigated. An AUC was calculated for each severity of PACG to evaluate the discriminatory accuracy of androstenedione in distinguishing between mild, moderate, and severe cases. The results of ROC analysis indicated AUC values ranging from 0.75 to 0.95 in discovery set 1 (Figure S10A, Table S8) and 0.94 to 0.99 in discovery set 2 (Figure S10B, Table S8) when using KNN machine learning algorithms. The validity of androstenedione in distinguishing the severity of PACG was demonstrated through consistent performance in both validation phase 1 (Figure S10C, Table S8, AUC of 0.64-0.97) and validation phase 2 (Figure S10D, Table S8, AUC of 0.98-1.0). Additionally, when mild and moderate cases were combined, the AUC for mild and moderate versus severe was 0.94 (95% CI 0.89 to 0.99) in discovery set 1 (Figure S9F, Table S8), 0.93 (95% CI 0.88 to 0.98) in discovery set 2 (Figure S9G, Table S8), 0.92 (95% CI 0.85 to 0.99) in validation phase 1 (Figure S9H, Table S8), and 0.98 (95% CI 0.96 to 1.0) in validation phase 2 (Figure S9I, Table S8).

### 3.7 Clinical value of androstenedione in patients with PACG

The lack of specificity of serum biomarkers remains a significant obstacle to the clinical application of such markers. To investigate temporal changes in androstenedione levels during the initial diagnosis and post-treatment period, we conducted a random analysis of nine pairs of blood samples from patients taken before and three months after surgical treatment (Figure 6A). Our findings indicate a significant decrease in androstenedione levels in the post-treatment serum of the nine patients compared to the pre-treatment serum (P = 0.021, Figure 6B).

**Figure 6.**
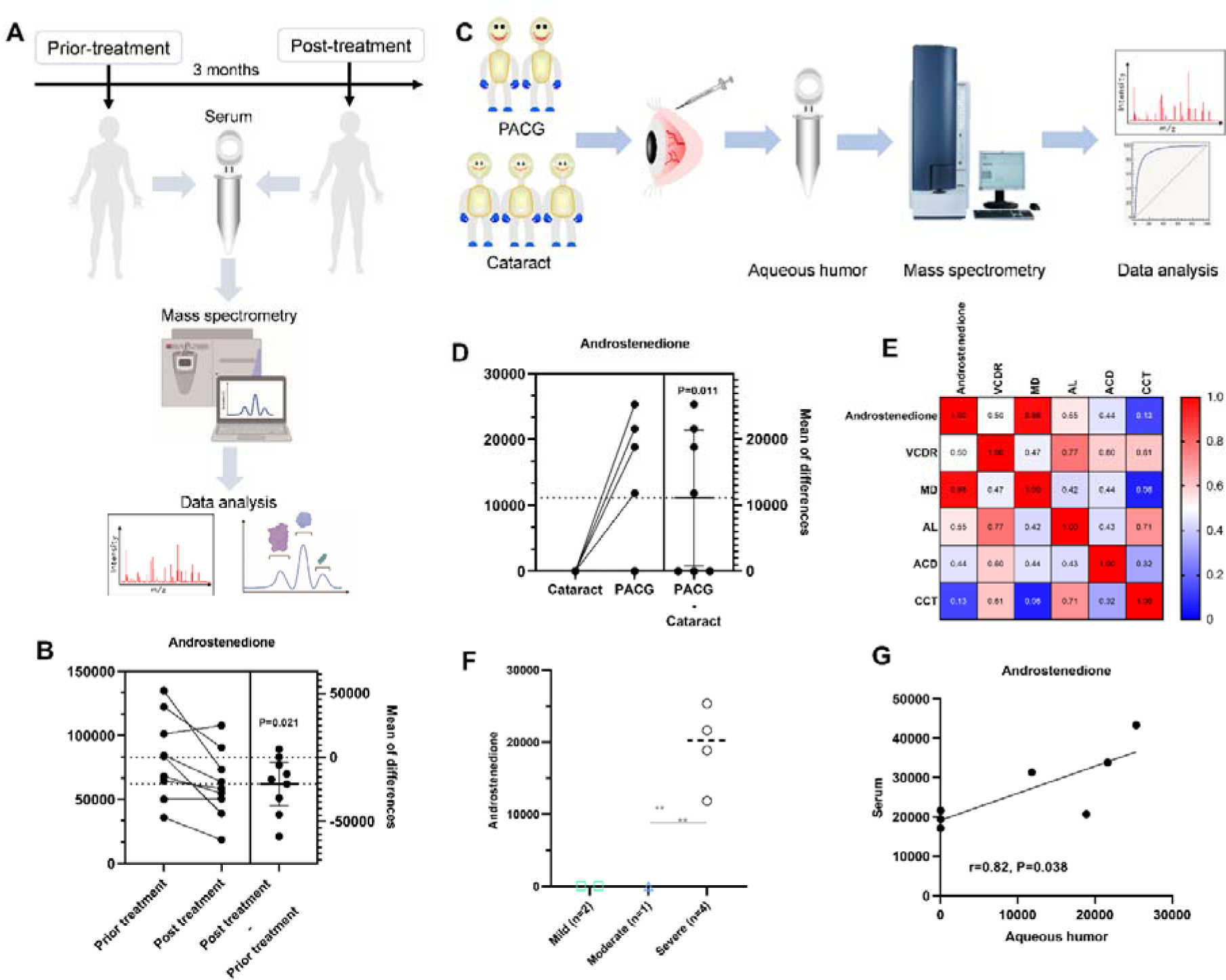
Specificity of circulating androstenedione in patients with primary angle closure glaucoma (PACG) in supplemental phase. A: Sampling scheme and workflow to investigate the temporal changes in androstenedione levels. B: Differential level of serum androstenedione between patients with PACG before and three months after treatment (Unit for y-axis is peak areas). C: Sampling scheme and workflow to determine whether aqueous humor levels of androstenedione were high in patients with PACG. D: The aqueous humor level of androstenedione between PACG and cataract (Unit for y-axis is peak areas). E: Heatmap of correlation analysis between ocular clinical characteristics and aqueous humor level of androstenedione. F: Comparison means aqueous humor levels of androstenedione between mild, moderate, and severe PACG (Unit for y-axis is peak areas). G: 7 paired serum-aqueous humor samples from the same PACG patients were included (Unit for y and x-axis is peak areas). A significant correlation between serum and aqueous humor levels of androstenedione was observed. Kruskal-Wallis test and one-way ANOVA was used. *P<0.05; **: P<0.001.

A case-control study (PACG=7, Cataract=11) (Figure 6C) was conducted to investigate the potential elevation of aqueous humor levels of androstenedione in patients. The findings revealed a statistically significant increase (P=0.011) in the levels of androstenedione in the aqueous humor of patients with PACG compared to those with cataracts (Figure 6D). Additionally, a significantly positive correlation between MD and aqueous humor levels of androstenedione was observed (Figure 6E, r=0.98, P<0.001). The mean aqueous humor levels of androstenedione were found to be significantly higher (P<0.001) in the severe PACG group compared to the moderate and mild PACG groups (Figure 6F). Subsequently, an examination was conducted on seven paired serum-aqueous humor samples obtained from identical PACG patients to ascertain the presence of a consistent pattern within the same individuals. A statistically significant correlation was observed between the levels of androstenedione in serum and aqueous humor (r=0.82, P=0.038) (Figure 6G).

### 3.8 Calibration ability of androstenedione on discovery phase and validation phase

Figure S11 displays the calibration plots for both the discovery and validation phases, which effectively validate the calibration performance of serum androstenedione for probability. The plots demonstrate a high level of agreement between predicted and observed values in both discovery set 1 (Figure S11A) and discovery set 2 (Figure S11B). The Hosmer-Lemeshow test yielded a nonsignificant statistic in the discovery set 1 (χ^2^=0, P=1) and discovery set 2 (χ^2^=6.16, P=0.10), indicating no departure from a perfect fit. Validation phase 1 (Figure S11C, χ^2^=5.14, P=0.14) and validation phase 2 (Figure S11D, χ^2^=1.25, P=0.26) resulted in similar performance.

### 3.9 Androstenedione could predict VF progression in patients with PACG

This study comprised 97 participants diagnosed with PACG, selected based on the screening criteria and followed up for a period of 24 months. In cases of bilateral PACG, one eye was selected at random. Of the total participants, 44 (45.36%) exhibited glaucoma progression, as evidenced by visual field (VF) loss. The demographic and ocular features of the VF progressing and non-progressing groups at baseline are presented in Table S9.

The patients in the progression group exhibited a statistically significant increase (P<0.001) in the mean serum levels of androstenedione compared to those in the non-progressing group (refer to Table S9). Furthermore, the multivariate Cox analysis revealed that the baseline levels of androstenedione (HR=2.71, 95% CI=1.20–6.10, P=0.017) were significantly associated with glaucoma progression, as determined by the VF loss results (Table 2). Figure 7 displays the Kaplan-Meier survival curves. The results of the survival analysis revealed a statistically significant increase in the proportion of patients with elevated androstenedione levels who experienced VF progression in PACG (log-rank test, P<0.001, Figure 7A). Comparable findings were observed in both the female (log-rank test, P=0.0042, Figure 7B) and male (log-rank test, P=0.0014, Figure 7C) subgroups.

**Figure 7.**
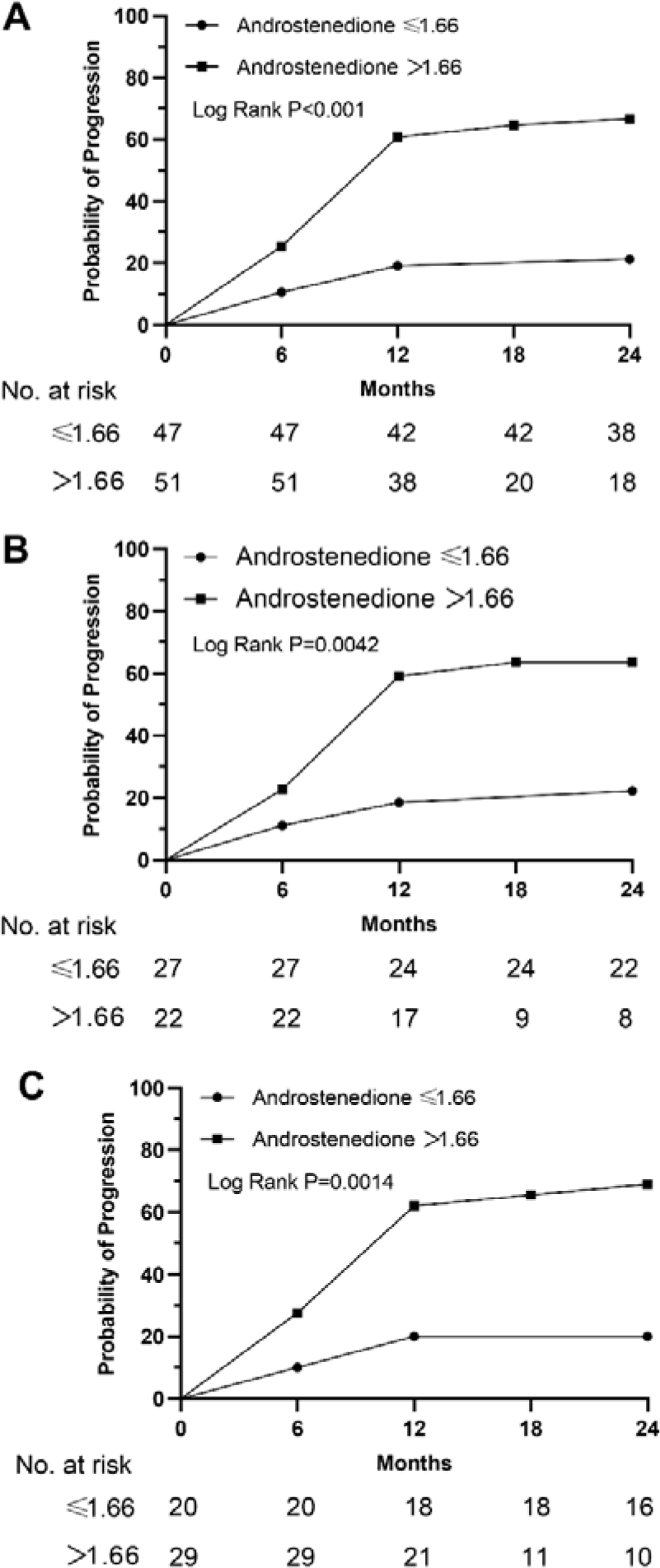
Kaplan-Meier curves stratified by the men value in terms of androstenedione. A: male+female; B: Female; C: Male. We categorized study participants into 2 groups based on their mean levels of androstenedione.

**Table 2.** Cox Proportional Hazards Regression Analysis to Assess the Value of Androstenedione Associated with Progression of PACG.

|  | Univariate |  | Multivariate* |  |
| --- | --- | --- | --- | --- |
|  | P | HR (95%CI) | P | HR (95%CI) |
| Age | **0.008** | **0.97 (0.95 to 0.99)** | 0.22 | 0.98 (0.95 to 1.01) |
| Sex | 0.87 | 0.95 (0.53 to 1.72) | 0.73 | 1.15 (0.53 to 2.52) |
| IOP | 0.59 | 1.007 (0.98 to 1.04) | 0.75 | 0.99 (0.96 to 1.03) |
| VCDR | 0.17 | 0.40 (0.11 to 1.46) | 0.16 | 0.37 (0.092 to 1.46) |
| CCT | 0.77 | 1.001 (0.10 to 1.007) | 0.90 | 1.000 (0.99 to 1.007) |
| ACD | **0.03** | **1.63 (1.05 to 2.52)** | 0.91 | 1.041 (0.51 to 2.13) |
| AL | **0.005** | **1.24 (1.07 to 1.44)** | 0.35 | 1.12 (0.89 to 1.41) |
| MD | 0.13 | 1.04 (0.99 to 1.08) | 0.075 | 1.047 (0.10 to 1.10) |
| Androstenedione | **<0.001** | **3.73 (1.84 to 7.57)** | **0.017** | **2.71 (1.20 to 6.10)** |

IOP = intraocular pressure, VCDR = vertical cup-to-disc ratio, AL = axial length, CCT = central corneal thickness, ACD = anterior chamber depth, MD = mean deviation, PACG=primary angle closure glaucoma.
\*adjusted for BMI, diabetes (yes=1, no=0), hypertension (yes=1, no=0), hypercholesterolemia (yes=1, no=0), smoking (yes=1, no=0), and drinking (yes=1, no=0).

## 4. Discussion

The delayed identification of PACG is a notable contributor to patients’ impaired vision and irreversible blindness. Regrettably, existing medical protocols do not advocate for the use of blood-based biomarkers to diagnose and prognosticate PACG. Consequently, it is imperative to develop novel, uncomplicated, and practicable approaches to improve the early detection of PACG and its predictive accuracy. To address the significant challenge at hand, the present study conducted a cross-sectional and prospective cohort investigation utilizing high-throughput, widely-targeted metabolomics and targeted chemiluminescence immunoassay on blood samples obtained from a substantial number of patients diagnosed with PACG and healthy individuals. The findings of this study reveal, for the first time, that androstenedione exhibits a reasonable level of precision (AUC, 0.85-1.0) in distinguishing between PACG and control groups in blood samples. Additionally, the baseline levels of androstenedione may serve as a valuable predictor of glaucomatous visual field progression.

Some metabolites have been proposed to have a potential role in discriminating PACG from normal, but validation studies in larger cohorts still need to be completed. Rong et al. ^14^ conducted a case-control (PACG=38, normal=48) study using gas chromatography-mass spectrometry and reported that palmitoleic acid, linoleic acid, γ-linolenic acid, and arachidonic acid were identified as essential metabolites associated with PACG, but diagnostic accuracy is unknown. In our study, these metabolites were also detected, but diagnosis accuracy (AUC<0.7) was limited. Qin et al.^24^ measured plasma twenty-two free fatty acids (FFA) and six lipid classes using metabolomics analysis, shown that docosahexaenoic acid (DHA) and total saturated fatty acids may be screening indices (AUC, 0.82-0.85) for PACG patients but lack validation set to confirm the results. DHA (AUC, 0.74-0.85), FFA (22:6) (AUC, 0.74-0.85), and FFA (18:4) (AUC, 0.79-1.0) was also shown a diagnose value for PACG patients in the discovery set of our study (Figure 3A), but the robustness and diagnose accuracy was weaker than androstenedione (AUC, 0.85-1.0) (Table S10). The contribution of previous metabolites to PACG diagnosis was small, which prompted us to conduct a trial designed to develop biomarkers.

In this study, androstenedione levels achieved better diagnostic efficiency and robust calibration across the discovery and validation sets. The transformation of normal subjects into PACG is a chronic stepwise process. Given this, screening healthy individuals and those with a family history of the disease is vital. Another important finding of this research was that the level of serum androstenedione retained its diagnostic efficiency for distinguishing the severity of PACG. Furthermore, serum androstenedione levels incrementally increased from mild to moderate to severe PACG, suggesting that serum androstenedione levels may accurately reflect the progression/severity of PACG. Furthermore, androstenedione levels at baseline were a new biomarker for predicting glaucomatous VF progression. Hence, serum androstenedione levels may provide a new biomarker for early detection and monitoring/predicting PACG severity/progression.

In the supplemental phase, we asked whether the serum levels of androstenedione were significantly correlated with aqueous humor levels of androstenedione from the same PACG patients, which is an important criterion for application as a routine clinical biomarker and to explore the biological function in PACG. Fortunately, the levels of aqueous humor androstenedione were positively correlated (r=0.82, P=0.038) with serum levels. Meanwhile, the levels of aqueous humor androstenedione incrementally increased from mild-moderate to severe PACG. Furthermore, the levels of aqueous humor androstenedione remained higher in PACG than in cataracts. Together, these results confirm that androstenedione can serve as a novel biomarker for early detection and monitoring of the progression/severity of PACG.

In clinical practice, oral topical glaucoma medications and laser peripheral iridectomy were the primary therapies used to treat PACG. However, the effects of these therapies are mainly estimated by measurement of intraocular pressure and optical coherence tomography. Thus, we have wondered whether, by dynamic detection, serum androstenedione levels could reflect the therapeutic effect of these therapies. In the supplemental phase (fourth phase), in a small cohort of 9 PACG patients treated with laser peripheral iridectomy or oral topical glaucoma medications, which showed a better prognosis, 7 of 9 exhibited decreased levels of serum androstenedione. These results shed new light on the monitor the effect of therapy; however, these should be verified in a larger cohort.

The main strength of our study is its robustness. We conducted a 5-phase study (discovery set [discovery set 1, the discovery set 2], validation phase 1, validation phase 2, supplemental phase, and cohort phase). We used large and well-characterized patients with adequate controls to confirm the results. Widely-targeted metabolomics and target chemiluminescence immunoassay methods provide high reliability of the metabolite. Androstenedione achieved better diagnostic accuracy across the discovery and validation sets, with AUC varying between 0.85 and 1.0. Interestingly, baseline androstenedione levels can predict glaucoma progression via VF loss results. Of note, the clinical practice of androstenedione in patients with PACG was analyzed by supplemental phase.

The metabolites identified by this study match PACG pathophysiological concepts. Pathway enrichment analysis demonstrated that these 32 significantly altered metabolites primarily belonged to 16 pathways (Figure S12). Among the top altered pathways, steroid hormone biosynthesis appears to be the critical node to high-match PACG pathophysiological concepts^25,26^: it connects with androstenedione. The molecular formula of androstenedione is shown in Figure S13. Steroid hormone biosynthesis appears to be a key node in the pathophysiological concept of highly matched PACG, but high enrichment is observed in metabolic pathways. Hormones play critical roles in various physiological functions, such as metabolism, immune responses, and inflammation regulation. In a related study on fatigue experienced during Androgen Deprivation Therapy (ADT), marked disparities were found in metabolite levels in the steroid hormone biosynthesis pathways, underscoring their significance in metabolic shifts. These insights highlight the intricate interconnection between steroid hormone biosynthesis and other metabolic pathways^27,28^. The sex hormones pathway is a vital component of steroid hormone biosynthesis (Figure S14A). Several studies have focused on the importance of 17β-estradiol in protecting the retinal ganglion cell layer and preserving visual function in clinical^29^ and basic^25^ research by anti-inflammatory effect^30^. Our previous studies suggested that decreased sex hormone concentrations in glaucoma lead to a hyperinflammatory state, leading to faster rates of VF damage^18,31^. Thus, we hypothesized that aromatase defects might lead to a decrease in estradiol levels and an increase in androstenedione, causing an inflammatory response and leading to retinal ganglion cell death in patient with glaucoma (Figure S14B). In addition, androstenedione converted to estrogens is catalyzed by the aromatase (Figure S14C). Furthermore, some of the metabolites might also be linked to PACG physiological processes that remain unclear, and future research in this field is needed. Sex hormones, including androstenedione, might be associated with glaucoma types such as POAG, normal-tension glaucoma, and pseudoexfoliation glaucoma^32^. The precise molecular mechanisms remain unclear. More research is essential to understand their relationship with different glaucoma types.

Furthermore, other studies have also found significant correlations between androstenedione and other diseases. For example, Javier et al.^33^ found that androstenedione can predict the progression of Frailty Syndrome in patients with localized breast cancer treated with aromatase inhibitors. Adriaansen et al.^34^ reported that diurnal salivary androstenedione levels in healthy volunteers for monitoring treatment efficacy of patients with congenital adrenal hyperplasia.

As far as our understanding goes, this is the initial investigation to methodically outline blood metabolites and scrutinize the diagnostic efficacy of a potential biomarker for PACG. Nevertheless, our study is not without its limitations: (1) The supplementary phase of our study was conducted with a restricted number of patients and healthy controls. Therefore, further investigations with larger sample sizes are necessary to confirm the diagnostic accuracy of serum androstenedione for the detection of primary angle-closure glaucoma (PACG). (2) Our study cohort exhibited uniform genetic and environmental traits, which may restrict the generalizability of our findings to populations with diverse ethnic or racial backgrounds. (3) Despite the matching of PACG patients and controls for age, gender, BMI, hypercholesterolemia, hypertension, diabetes, smoking, and drinking, it is possible that unidentified residual confounding factors could impact the observed metabolomic disparities. (4) Throughout the duration of the follow-up period, the majority of individuals received pharmacological treatment, and the study’s follow-up period was limited to a mere two years. Consequently, the outcomes may have been impacted by either behavioral adjustments prompted by patients’ cognizance of their medical condition or by any form of therapeutic intervention. (5) Despite our examination of the impact of hormone intake (including estrogen, progestagen, and anti-androgen), the potential influence of reproductive aging cannot be entirely dismissed. (6) Understanding the link between changes in androstendione levels and glaucoma severity might hinge on metabolic and anti-inflammatory pathways. However, the current study did not delve deeply into the mechanism verification. Further research is essential to comprehensively grasp the precise relationship between androstendione and the severity of glaucoma, as well as the mechanisms at play.

## 5. Conclusions

In conclusion, androstenedione has been identified and validated through the use of widely-targeted LC-MS or targeted chemiluminescence immunoassay, demonstrating its ability to effectively differentiate between healthy individuals and those with PACG. Additionally, baseline androstenedione levels may serve as a valuable predictor of glaucomatous VF progression. However, further clinical and basic studies are required to confirm the clinical utility of serum androstenedione for early-stage PACG diagnosis, as well as its potential for monitoring/predicting VF progression and elucidating the underlying mechanisms linking androstenedione to PACG.

## Declarations

### Ethics approval and consent to participate

This study was approved by the Ethics Committee of Eye and ENT Hospital of Fudan University (EENT-2015011) and was conducted under the Declaration of Helsinki. All participants provided written informed consent prior to their participation.

### Consent for publication

Not applicable.

### Availability of data and materials

The datasets used and/or analyzed during the current study are available from the corresponding author on reasonable request.

### Authors’ contributions

Shengjie Li, Shunxiang Gao and Wenjun Cao contributed to the study conception and design, data analysis, interpretation of the data, and drafting the manuscript. Yichao Qiu, Jun Ren, Zhendong Jiang, Yingzhu Li, Yunxiao Song, and Mingxi Shao contributed to the interpretation of the data and critical revision of the manuscript. Shengjie Li, Yichao Qiu, Jun Ren, Yingzhu Li, Yunxiao Song, Xinghuai Sun, Shunxiang Gao and Mingxi Shao contributed to the collection of the data. All authors read and approved the final manuscript.

## Data Availability

All data produced in the present study are available upon reasonable request to the authors

## Acknowledgements

Not applicable.

